# Production and performance assessment of a SARS-CoV-2 biomimetic in a verification program for pandemic readiness

**DOI:** 10.1101/2023.06.26.23291917

**Authors:** Edith Erika Machowski, Anna Esther Reyneke, Dean Evan Sher, Bavesh Davandra Kana

## Abstract

During the early stages of the Covid-19 pandemic in South Africa, one of many challenges included availability of control material proficiency testing programs. Control material utilising live SARS-CoV-2 or RNA extracted from cell culture was either biohazardous and costly, particularly in resource limited settings. Here, we report the development and application of a non-infectious SARS-CoV-2 biomimetic *Mycobacterium smegmatis* strain that mimics a positive result in the GeneXpert SARS-CoV-2 Xpert Xpress cartridge. Nucleotide sequences located in genes encoding the RNA-dependent RNA polymerase, the nucleocapsid and the envelope proteins were used. The resulting biomimetic was prepared as a Quality Control specimen and distributed to laboratories in South Africa for validation prior to testing of clinical specimens. Between April 2020 and December 2020, a total of 151 instruments were validated to bring Covid-19 mass testing online. These instruments capacitated the country to perform tests in 2532 modules. False negative or false positive findings reflected issues such as workflow/technician error or other related technical issues. This non-infectious, easily scalable control material became available within two months after the start of the pandemic in South Africa and represents a useful approach to consider for other diseases and future pandemics.

## Introduction

SARS-CoV-2, the causative agent of Covid-19, was first described in December 2019, subsequently progressing to a global pandemic (Figure 1). It was initially designated as 2019-nCoV but subsequently renamed (1-6). It is a *ß*-coronavirus with a single stranded RNA genome of 29 903 nucleotides (Strain Wuhan-Hu-1 accession number MN908947.3). Early during the pandemic, it became evident that effective control required fast and accurate diagnosis, as the inability to rapidly identify infected/diseased individuals fuelled further transmission. To detect SARS-CoV-2, the presence of its genome in clinical specimens needs to be established by means of nucleic acid amplification technologies (NAAT). Sequence information and assay methodology for early diagnostic assays were made publicly available to assist global health systems with implementing pandemic control measures (7).

**Figure 1.**
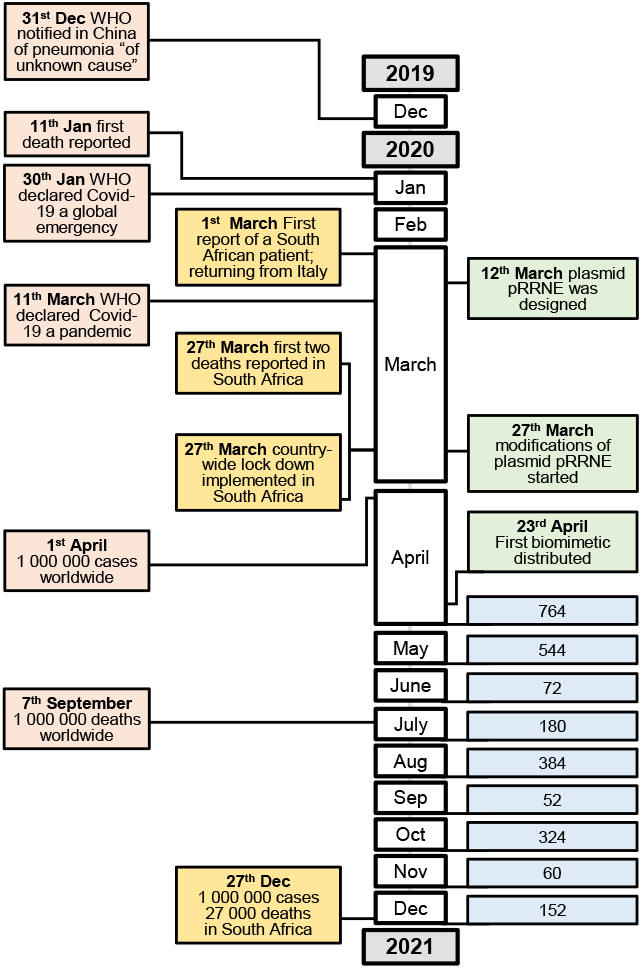
Timeline of the Covid Pandemic. The first column in pink indicates the worldwide progression of clinical cases and response from the World Health Organization. The second column in orange indicates the progression of clinical cases in South Africa and the response from the South African National Department of Health. The third column shows the dates from December 2019 to December 2020 (not to scale). The fourth column shows the progress of plasmid and strain development in green, and the number of GeneXpert modules verified in each month between April and December 2020 in blue. This illustrates the rapid development of biomimicry approaches to enable rollout of mass Covid-19 testing.

Global deployment of NAAT, particularly in resource limited settings, was difficult at the time due to constraints on tools, materials, trained technicians and laboratory resources (8). The lack of robust, consistent controls to verify the performance of emerging Covid-19 diagnostics was a significant barrier to mass testing. Positive clinical specimens were used, alternatively purified viral RNA served as a positive control, but this was problematic and costly due to its chemical instability, the need for cold chain storage and limited availability. In addition to using clinical specimens, deployment of RNA controls also required the capacity to culture live SARS-CoV-2, in bulk, using cell culture to yield sufficient amounts. Given the pathogenic nature of the virus, this approach was fraught with challenges.

We previously reported the use of a modified, non-infectious mycobacterial species for use in instrument verification and external quality assurance (EQA) in tuberculosis molecular diagnostics programs, which faced similar challenges with respect to biologically safe controls (9). These biomimetic strains were able to mimic positive tuberculosis diagnosis on two distinct platforms, with this approach also demonstrating utility for detection of a *Staphylococcus aureus* target sequence. Based on the sequence of NAAT targets available during 2020 at the time of the Covid-19 outbreak, we adapted this biomimetic approach to generate control material that was non-infectious and easily scalable in South Africa. Herein, we describe the validation procedure for SARS-CoV-2 diagnosis that was implemented in 2020 in South Africa, using a biomimetic strain of *Mycobacterium smegmatis* that yields the expected positive result in the GeneXpert SARS-CoV-2 Xpert Xpress cartridge. We anticipate that this approach can be adopted to rapidly scale diagnostics for other diseases and future pandemics.

## Materials and Methods

### Culture conditions

All strains and plasmids used and generated in this study are listed in Table 1. Strain *Escherichia coli* DH5α was grown at 37°C shaking in standard lysogeny broth (LB) or on Luria agar (LA), supplemented with 50 µg/ml of kanamycin (Kan) where applicable. *M. smegmatis* strains were grown at 37°C shaking in Middlebrook 7H9 liquid medium (Difco) supplemented with 0.085% NaCl, 0.2% glucose, 0.2% glycerol, and 0.05% Tween-80 or on Middlebrook 7H10 solid medium (Difco) supplemented with 0.085% NaCl, 0.2% glucose, and 0.5% glycerol. Kan was used at 25 µg/ml.

**Table 1.**
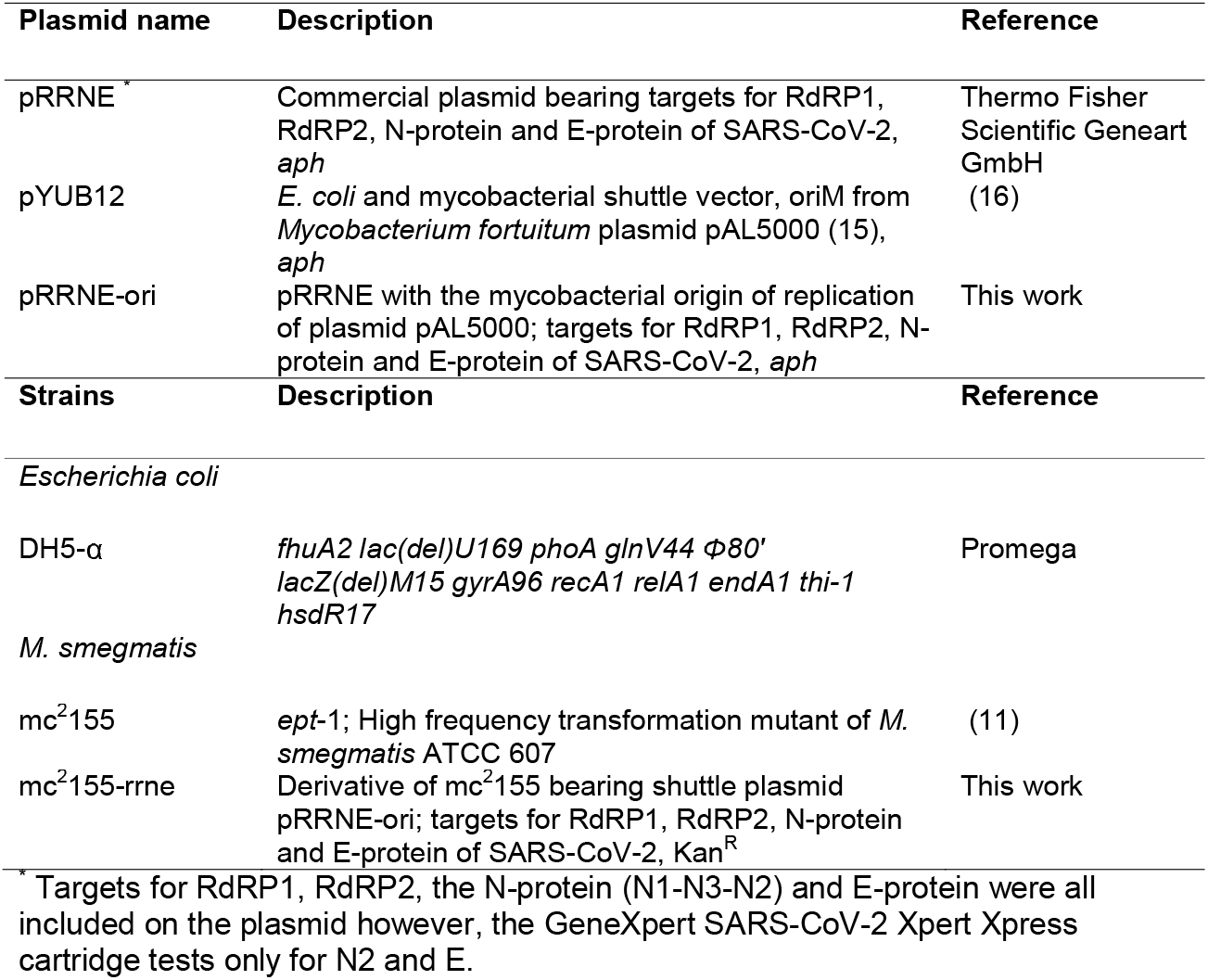
Plasmids and bacterial strains generated or used in this study.

### Design of shuttle vectors and biomimetic strains

The complete genomic organization of SARS-CoV-2 is shown in Figure 2A and selected target regions for generating a biomimetic are shown in Figure 2B. Plasmid pRRNE was purchased commercially and carried the target sequences for the regions in the SARS-CoV-2 RNA-dependent-RNA polymerase, (RdRP1, RdRP2), the N-protein (N1-N3-N2) and the E-protein (E). These sequences were made publicly available in January 2020 (10). pRRNE was introduced into *M. smegmatis*, in combination with a mycobacterial origin of replication by standard electroporation methodology (11), to yield strain mc^2^155-rrne as a positive quality control (QC) specimen.

**Figure 2.**
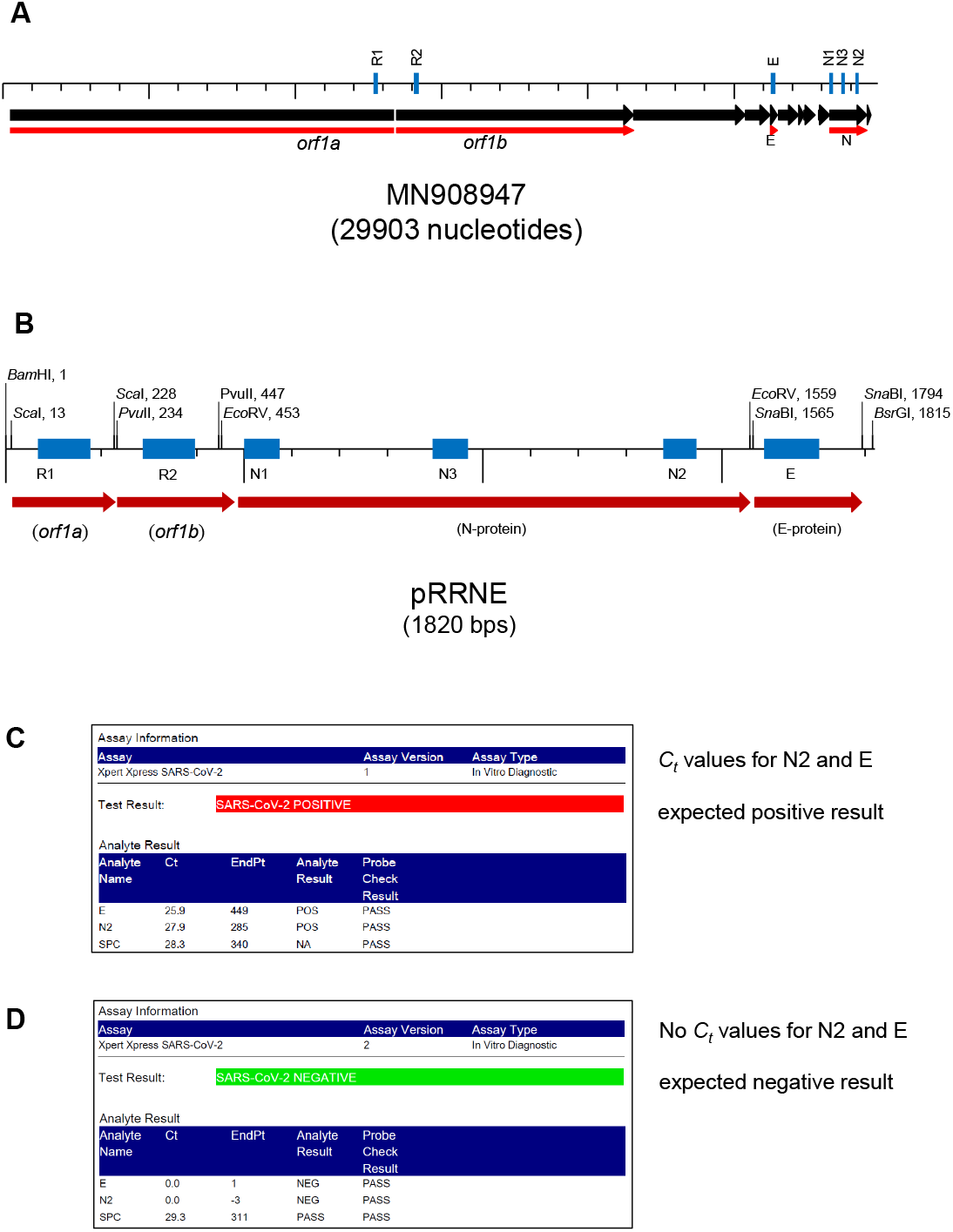
Nucleotide Sequence of SARS-CoV-2, Genbank Accession number MN908947 and representative positive and negative test results. **A**. Genomic sequence of the entire single-stranded RNA coding molecule. black, annotated genes; red, the genes in which target sequences are located; blue, the target locations published by Corman et al. (2020) **B**. The sequence designed and purchased for this study, based on the targets indicated in panel A. Unique restriction enzyme recognition sequences were included flanking each target. **C**. Positive result for the biomimetic strain mc^2^155-rrne. **D**. Negative result for a QC specimen.

### Distribution of QC Specimens

Verification panels contained two QC specimens each, one negative and one positive (Figure 3). One verification panel was distributed to each GeneXpert instrument in laboratories enrolled in the SmartSpot Quality SARS-CoV-2 verification program.

**Figure 3.**
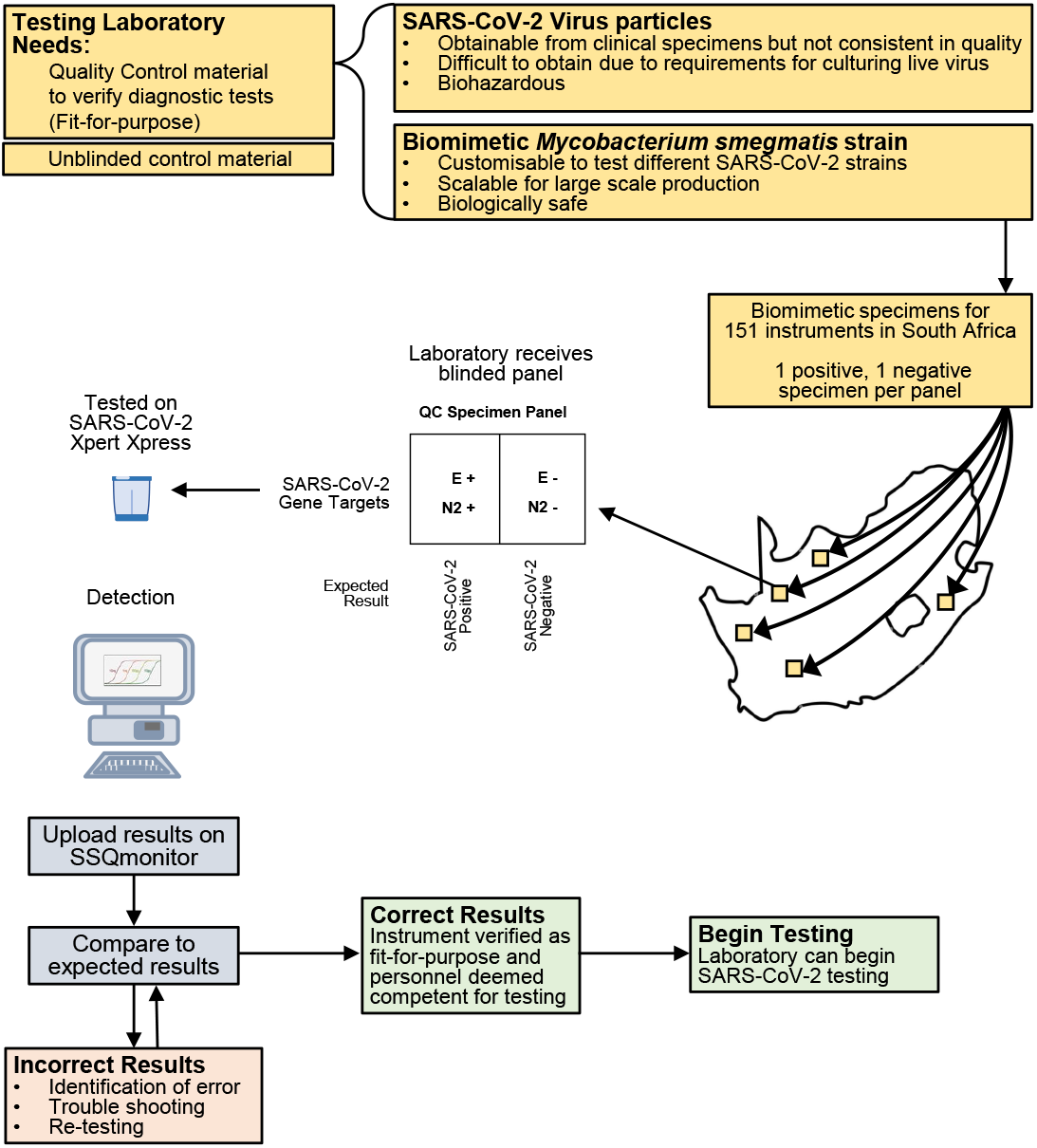
Distribution of QC specimens for module verification and Analysis of QC test results. Verification panels were shipped to laboratories on demand. Tests were performed on GeneXpert modules for detection in the SARS-CoV-2 Xpert Xpress cartridge. Incorrect results required assessment and troubleshooting to ensure that the source of errors was identified and corrected before bringing a laboratory online.

### Data Collection and Analysis

Each affiliated laboratory exported their results from their GeneXpert instrument software and uploaded their results to SmartSpot Quality via its online result reporting tool, www.SSQmonitor.com (Figure 3). The results were auto-analysed on SSQmonitor, and where full scores were achieved, QC Reports were released in real-time. Where results were not concordant, reports were held back for review and analysed in batches prior to report finalization. Where warranted, *C*_*t*_ values were analysed to facilitate accurate result scoring and to identify and address potential root causes for incorrect results.

## Results

### Test Result of SARS-CoV-2 Xpert Xpress analysis

Biomimetic strain mc^2^155-rrne was validated in the SARS-CoV-2 Xpert Xpress cartridge prior to launching the control in testing laboratories. It yielded the expected result of “positive”, (Figure 2C) based on the presence of C_*t*_ values for the E and N2 targets. In their absence the negative QC specimen yielded the expected result of “negative”, without C_*t*_ values (Figure 2D).

### Distribution of QC Panels

Verification Panels were shipped to laboratories to bring GeneXpert modules online for testing, where one module is the unit in a GeneXpert machine that can test one cartridge at a time (Figure 3). The large demand for SARS-CoV-2 testing and reagents – together with lockdown regulations – limited access to resources, which were accordingly prioritized for patient testing. To minimize the number of cartridges required to verify the SARS-CoV-2 assay for validation in any given laboratory, machines that were already enrolled for standard diagnosis of other diseases such as tuberculosis, and were thus already verified with other QC specimens, only received one positive and one negative specimen. Sixty five laboratories across South Africa were enrolled in the verification program with a total of 151 instruments, which effectively comprised 2532 modules. A total of 702 QC specimens were distributed, of which 349 positive specimens and 350 negative specimens were reported on (Table 2). Discrepancies in numbers of specimens submitted indicate that some laboratories did not test both specimens supplied within the verification panel. This was attributed to either reagent shortages, human error in processing the specimen/s and/or human error in submitting both results.

**Table 2.** Test Result of Xpert Xpress SARS-CoV-2 analysis.

|  | Number of specimens Tested | Correct (%) | Incorrect (%) |  |  |  |
| --- | --- | --- | --- | --- | --- | --- |
|  |  |  | Positive | Presumptive* | Negative | Error |
| **Positive QC specimen** | 349 | 345 (98.9) | - | 0 (0.0) | 3 (0.9) | 1 (0.3) |
| **Negative QC specimen** | 350 | 343 (98.0) | 6 (1.7) | 1 (0.3) | - | 0 (0.0) |

\* A presumptive positive result indicates a specimen that only flags positive for the E gene, which is indicative of any $\beta$ -coronavirus, not only SARS-CoV-2 {Coronaviridae Study Group of the International Committee on Taxonomy of, 2020 #1}

### Reliability of the QC Specimens

The verification panels yielded 98.9% (345/349) correct positive results and 98.0% (343/350) correct negative results, with an overall concordance of 98.9% (Table 2). The incorrect results submitted for the negative QC specimens were as follows: 1.7% (6/350) reported as positive and 0.3% (1/350) reported as presumptive positive (A C_*t*_ value was obtained for analyte E and no C_*t*_ value for N2). The incorrect results submitted for the positive QC specimens were: 0.9% (3/349) reported as negative and 0.3% (1/349) reported as an instrument error.

### Assessment of laboratory competency

To assess the proficiency of testing facilities, a GeneXpert system was deemed fit-for-purpose provided that all modules that were tested within the panel passed verification. If any module did not pass verification, a repeat specimen was tested.

## Discussion

In this work, we demonstrate the utility of using a non-infectious biomimetic to generate biologically safe Covid-19 diagnostic controls, and integrating this into verification of a molecular diagnostic assay. The resulting biomimetics were robust and easily scalable as QC material for laboratories across an entire country. This approach enabled activation of 2532 GeneXpert modules for immediate SARS-CoV-2 mass testing. The application of this technology for SARS-CoV-2 can be applied to other diagnostic platforms, based on NAAT approaches. However, conditions that ensure lysis of the bacterial biomimetic cell wall may require some optimization on other platforms. Given the robust results obtained in this study, this issue did not emerge in the GeneXpert platform.

The GeneXpert cartridges are supplied with all components in stable format at room temperature (2 -28 °C), including the required enzymes, and an internal cartridge control that assures lysis and thermocycling perform adequately. In the field, SARS-CoV-2 Xpert Xpress performed well in side-by-side comparisons with other diagnostic platforms to detect the virus on clinical samples (12, 13). This confirms that both the reverse transcription and PCR steps are reproducible and reliable. Under this assumption, the *C*_*t*_ values and results obtained from the biomimetics can be considered a valid proxy for the identification of SARS-CoV-2 genome sequence. However, it should be noted that these biomimetic controls are DNA-based and as such, do not control for proficiency of the reverse transcription step.

A critical factor in the ease of implementation in the roll-out of diagnostic capability in South Africa was that GeneXpert technology already has a large presence due to its extensive use in tuberculosis diagnosis (Figure 3, (14)). For instrument verification we have shown that the results obtained from the biomimetic *M. smegmatis* SARS-CoV-2 QC specimens reliably mimic those expected from corresponding clinical samples, based on the presence or absence of the targets included in the material. This approach can be easily adapted for emerging pathogens and future pandemics and negates the requirement to handle large amounts of infectious specimens in proficiency testing programs. Given the increased regulation on expanding pathogens in laboratories, our approach hold promise for bolstering expanded rollout of molecular diagnostics.

## Author contributions

E.E.M. and B.D.K. conceived the study and designed the biomimetic strain. E.E.M. performed laboratory procedures to generate mc^2^155-rrne. A.E.R. and D.E.S. produced QC specimens from strain mc^2^155-rrne and led the distribution of specimens, data collection and data analysis of the study. All authors contributed to compiling the manuscript.

## Data Availability

All data produced in the present work are contained in the manuscript

## References

1. Coronaviridae Study Group of the International Committee on Taxonomy of V. The species Severe acute respiratory syndrome-related coronavirus: classifying 2019-nCoV and naming it SARS-CoV-2. Nat Microbiol. 2020;5(4):536–44.

2. WHO. Surveillance case definitions for human infection with novel coronavirus (nCoV): interim guidance, 11 January 2020.: World Health Organization; 2020.

3. Ren LL, Wang YM, Wu ZQ, Xiang ZC, Guo L, Xu T, et al. Identification of a novel coronavirus causing severe pneumonia in human: a descriptive study. Chin Med J (Engl). 2020;133(9):1015–24.

4. Wu F, Zhao S, Yu B, Chen YM, Wang W, Song ZG, et al. A new coronavirus associated with human respiratory disease in China. Nature. 2020;579(7798):265–9.

5. Wu F, Zhao S, Yu B, Chen YM, Wang W, Song ZG, et al. Author Correction: A new coronavirus associated with human respiratory disease in China. Nature. 2020;580(7803):E7.

6. Zhu N, Zhang D, Wang W, Li X, Yang B, Song J, et al. A Novel Coronavirus from Patients with Pneumonia in China, 2019. N Engl J Med. 2020;382(8):727–33.

7. WHO. Molecular assays to diagnose COVID-192020.

8. Reusken C, Broberg EK, Haagmans B, Meijer A, Corman VM, Papa A, et al. Laboratory readiness and response for novel coronavirus (2019-nCoV) in expert laboratories in 30 EU/EEA countries, January 2020. Euro Surveill. 2020;25(6).

9. Machowski EE, Kana BD. Genetic Mimetics of Mycobacterium tuberculosis and Methicillin-Resistant Staphylococcus aureus as Verification Standards for Molecular Diagnostics. J Clin Microbiol. 2017;55(12):3384–94.

10. Corman VM, Landt O, Kaiser M, Molenkamp R, Meijer A, Chu DK, et al. Detection of 2019 novel coronavirus (2019-nCoV) by real-time RT-PCR. Euro Surveill. 2020;25(3).

11. Snapper SB, Melton RE, Mustafa S, Kieser T, Jacobs WR, Jr. Isolation and characterization of efficient plasmid transformation mutants of Mycobacterium smegmatis. Mol Microbiol. 1990;4(11):1911–9.

12. Loeffelholz MJ, Alland D, Butler-Wu SM, Pandey U, Perno CF, Nava A, et al. Multicenter Evaluation of the Cepheid Xpert Xpress SARS-CoV-2 Test. J Clin Microbiol. 2020;58(8).

13. Cao XJ, Fang KY, Li YP, Zhou J, Guo XG. The Diagnostic Accuracy of Xpert Xpress to SARS-CoV-2: A systematic review. J Virol Methods. 2022;301:114460.

14. Scott LE, McCarthy K, Gous N, Nduna M, Van Rie A, Sanne I, et al. Comparison of Xpert MTB/RIF with other nucleic acid technologies for diagnosing pulmonary tuberculosis in a high HIV prevalence setting: a prospective study. PLoS Med. 2011;8(7):e1001061.

15. Rauzier J, Moniz-Pereira J, Gicquel-Sanzey B. Complete nucleotide sequence of pAL5000, a plasmid from Mycobacterium fortuitum. Gene. 1988;71(2):315–21.

16. Snapper SB, Lugosi L, Jekkel A, Melton RE, Kieser T, Bloom BR, et al. Lysogeny and transformation in mycobacteria: stable expression of foreign genes. Proc Natl Acad Sci U S A. 1988;85(18):6987–91.

